# Expert-level pediatric brain tumor segmentation in a limited data scenario with stepwise transfer learning

**DOI:** 10.1101/2023.06.29.23292048

**Authors:** Aidan Boyd, Zezhong Ye, Sanjay Prabhu, Michael C. Tjong, Yining Zha, Anna Zapaishchykova, Sridhar Vajapeyam, Hasaan Hayat, Rishi Chopra, Kevin X. Liu, Ali Nabavidazeh, Adam Resnick, Sabine Mueller, Daphne Haas-Kogan, Hugo J.W.L. Aerts, Tina Poussaint, Benjamin H. Kann

**Affiliations:** Artificial Intelligence in Medicine (AIM) Program, Mass General Brigham, Harvard Medical School, Boston, MA; Department of Radiation Oncology, Dana-Farber Cancer Institute and Brigham and Women’s Hospital, Harvard Medical School, Boston, MA; Department of Radiology, Boston Children’s Hospital, Harvard Medical School, Boston, MA; Center for Data-Driven Discovery in Biomedicine (D3b), Children’s Hospital of Philadelphia, Philadelphia, PA; Department of Radiology, Perelman School of Medicine, University of Pennsylvania, Philadelphia, PA; Department of Neurosurgery, Children’s Hospital of Philadelphia, Philadelphia, PA; Department of Neurology, University of California San Francisco, San Francisco, California; Department of Pediatrics, University of California San Francisco, San Francisco, California; Department of Neurological Surgery, University of California San Francisco, San Francisco, California; Department of Radiology, Brigham and Women’s Hospital, Dana-Farber Cancer Institute, Harvard Medical School, Boston, MA, USA; Radiology and Nuclear Medicine, CARIM & GROW, Maastricht University, Maastricht, the Netherlands

**Keywords:** MRI, Pediatric Low-Grade Glioma, Automatic Tumor Segmentation, nnUNet, Deep Neural Network

## Abstract

**Purpose:** Artificial intelligence (AI)-automated tumor delineation for pediatric gliomas would enable real-time volumetric evaluation to support diagnosis, treatment response assessment, and clinical decision-making. Auto-segmentation algorithms for pediatric tumors are rare, due to limited data availability, and algorithms have yet to demonstrate clinical translation.

**Methods:** We leveraged two datasets from a national brain tumor consortium (n=184) and a pediatric cancer center (n=100) to develop, externally validate, and clinically benchmark deep learning neural networks for pediatric low-grade glioma (pLGG) segmentation using a novel in-domain, stepwise transfer learning approach. The best model [via Dice similarity coefficient (DSC)] was externally validated and subject to randomized, blinded evaluation by three expert clinicians wherein clinicians assessed clinical acceptability of expert- and AI-generated segmentations via 10-point Likert scales and Turing tests.

**Results:** The best AI model utilized in-domain, stepwise transfer learning (median DSC: 0.877 [IQR 0.715-0.914]) versus baseline model (median DSC 0.812 [IQR 0.559-0.888]; *p*<0.05). On external testing (n=60), the AI model yielded accuracy comparable to inter-expert agreement (median DSC: 0.834 [IQR 0.726-0.901] vs. 0.861 [IQR 0.795-0.905], *p*=0.13). On clinical benchmarking (n=100 scans, 300 segmentations from 3 experts), the experts rated the AI model higher on average compared to other experts (median Likert rating: 9 [IQR 7-9]) vs. 7 [IQR 7-9], *p*<0.05 for each). Additionally, the AI segmentations had significantly higher (*p*<0.05) overall acceptability compared to experts on average (80.2% vs. 65.4%). Experts correctly predicted the origins of AI segmentations in an average of 26.0% of cases.

**Conclusions:** Stepwise transfer learning enabled expert-level, automated pediatric brain tumor auto-segmentation and volumetric measurement with a high level of clinical acceptability. This approach may enable development and translation of AI imaging segmentation algorithms in limited data scenarios.

**Summary:** Authors proposed and utilized a novel stepwise transfer learning approach to develop and externally validate a deep learning auto-segmentation model for pediatric low-grade glioma whose performance and clinical acceptability were on par with pediatric neuroradiologists and radiation oncologists.

**Key Points:**

- There are limited imaging data available to train deep learning tumor segmentation for pediatric brain tumors, and adult-centric models generalize poorly in the pediatric setting.
- Stepwise transfer learning demonstrated gains in deep learning segmentation performance (Dice score: 0.877 [IQR 0.715-0.914]) compared to other methodologies and yielded segmentation accuracy comparable to human experts on external validation.
- On blinded clinical acceptability testing, the model received higher average Likert score rating and clinical acceptability compared to other experts (*Transfer-Encoder* model vs. average expert: 80.2% vs. 65.4%)
- Turing tests showed uniformly low ability of experts’ ability to correctly identify the origins of *Transfer-Encoder* model segmentations as AI-generated versus human-generated (mean accuracy: 26%).

## Introduction

Pediatric low-grade gliomas (pLGGs) are the most common type of brain tumors in children, accounting for approximately 30% of all pediatric brain tumors (1). pLGGs are heterogeneous in their molecular underpinnings, natural history, and aggressiveness, and therapies carry significant risks, thus making management decisions challenging (2–4). Optimal risk-stratification, response assessment, and surveillance for pLGG hinge on the ability to accurately localize and characterize brain tumors on magnetic resonance imaging (MRI) scans, which, in turn, relies on accurate tumor segmentation. Compared to adult gliomas, manual segmentation of pLGG carries distinct challenges, requiring expertise, resources, and time, thus limiting its clinical efficiency (5).

Given the utility of accurate segmentation and the inherent limitations of manual segmentation, there has been interest in developing auto-segmentation tools for pediatric brain tumors (6–8). The progress in medical imaging techniques and computational methods have led to the development of various approaches for brain tumor segmentation, including machine learning-based methods, deep learning-based methods, and hybrid approaches (8–10). Recently, deep learning has emerged as a powerful tool in medical imaging, offering solutions to diverse clinical challenges (11–15). Auto-segmentation based on deep learning is thus a promissing approach for accurate and efficient brain tumor segmentation, including pLGGs (6,16,17), though distinct challenges remain. In particular, pLGGs are relatively rare tumors and there are no publicly available datasets for training models. Most brain tumor segmentation algorithms have been developed for adult glioma, which are much more common and have large volumes of public and institutional data for training (18,19). In contrast, there has been only limited study of dedicated pediatric glioma segmentation - with a paucity of pLGG-specific models that rely on small, single-institution datasets that have not been externally validated nor subjected to clinical testing (16,17). Human clinical evaluation of segmentation models is essential to benchmark performance to experts and determine their true level of performance and potential for clinical translation.

There are numerous proposed ways to improve deep learning performance in limited-data settings (20). Aside from traditional techniques like data augmentation, ensembling, regularization, recently, advances have been made in knowledge transfer-learning (21) and self- (22) and semi-supervision (23). These have shown promise in improving medical image analysis algorithms, though can be challenging to implement. Pediatric brain tumors represent an ideally situated setting for which to apply these techniques, given the relative scarcity of data. Here, we aim to bridge the translational gap for pediatric brain tumor segmentation algorithms and achieve clinically acceptable performance despite a limited-data scenario with several innovations: 1) implementation of in-domain stepwise transfer learning; 2) aggregation of the largest pLGG imaging database to-date, and 3) performing blinded human acceptability testing.

## MATERIALS AND METHODS

### Study Design and Datasets

This study was conducted in accordance with the Declaration of Helsinki guidelines and following the approval of the Dana-Farber/Boston Children’s/Harvard Cancer Center Institutional Review Board (IRB). Waiver of consent was obtained from IRB prior to research initiation due to public datasets or retrospective study. Data from one high-volume academic institution and one national consortium from 2001 through 2015 were included. The Children’s Brain Tumor Network (CBTN) dataset consists of one de-identified patient cohort (n=187). The Boston Children’s Hospital (BCH) dataset includes one patient cohort (n=100). These subsets represent all scans that passed initial quality control of DICOM metadata. Patient inclusion criteria were the following: 1) 0–18 years of age, 2) histopathologically confirmed pLGG, and 3) availability of preoperative brain MR imaging with a T2-weighted (T2W) imaging sequence. Spinal cord tumors were not considered for this study. Scans with spine tumors (n=12) and undetectable lesions (n=3) were excluded from analyses. Publicly available data from the Brain Tumor Segmentation (BraTS) 2021 competition was acquired from http://braintumorsegmentation.org (24–26). The BraTS competition data represents 1,251 adult glioma multiparametric MR scans along with expert-generated segmentation masks for T1-weighted (T1W), T2W, fluid attenuated inversion recovery (FLAIR), and contrast enhancement generated by radiologists (24,25). Full data description for BraTS is available here: http://braintumorsegmentation.org.

### MR Imaging Acquisition and Parameters

Patients from the CBTN cohort underwent brain MR imaging at 1.5T or 3T Siemens MR scanners (Table S1). Sequences acquired included 2D axial T2-weighted turbo spin-echo (TR/TE, 1000– 7300/80–530 ms; 0.5- to 5-mm section thickness), 3D axial or sagittal pre-contrast, and 3D axial gadolinium-based contrast agent–enhanced T1-weighted turbo or fast-field echo. Patients from the BCH cohort underwent brain MR imaging at 1.5T or 3T from various MRI vendors (Table S2). MRIs were performed using the brain tumor protocol of the institution, which included 2D axial T2-weighted fast spin-echo (TR/TE, 2600–12,000/11–120 ms; 2- to 5.5-mm section thickness), 2D axial or sagittal pre-contrast T1-weighted spin-echo, 2D axial T2 FLAIR and 2D axial gadolinium-based contrast agent–enhanced T1-weighted spin-echo sequences (Table S1-S2; Fig. S1-S4). All MR imaging data were extracted from the respective PACS and were de-identified for further analyses. Given that many pLGGs do not enhance with intravenous contrast, are hypointense on T1, and are hyperintense on T2-weighted sequences, we chose to develop a T2-weighted segmentation algorithm with a primary use case of volumetric tumor monitoring.

### MR Image Preprocessing

MRI images were converted from DICOM format to NIFTI format via rasterization packages utilizing dcm2nii package (https://www.nitrc.org/projects/dcm2nii) in Python v3.8. N4 bias filed correction was adopted to correct the low-frequency intensity non-uniformity present in MRI images using SimpleITK (SITK) in Python v3.8. All scans were resampled to 1ξ1ξ3 mm^3^ voxel size using linear interpolation via SITK. After interpolation, the MR scans were co-registered using a rigid registration step with SITK. Lastly, a brain extraction step was performed for all the scans using HD-BET package in Python v3.8 (27). Preprocessing scripts are found here: https://github.com/BoydAidan/nnUNet_pLGG.

### MR image review and segmentation

For model development and initial training, tumors in all T2w scans within the CBTN and 60 scans within the BCH cohort were initially segmented by a board-certified radiation oncologist (experience: 8 years) to serve as primary ground truth segmentations. Annotators were instructed to segment all areas of T2 signal abnormality concerning for tumor involvement, including areas of peritumoral T2 hyperintensity if suspicious for tumor involvement. Segmentations were performed and saved in NIFTI format using ITK-SNAP v4.0 (http://www.itksnap.org) in 3D utilizing axial, sagittal, and coronal views (Fig. S5).

### Deep learning approach: stepwise, in-domain transfer learning

For this work, the nnUNet architecture (28) which is a deep learning-based segmentation approach that configures itself completely automatically, was chosen for the deep learning framework. nnUNet includes auto-configuration for preprocessing, network architecture selection, model training and post-processing and can be applied to any task. nnUNet performs well out of the box on many medical imaging tasks, and it has been specifically shown to produce strong results for brain tumor segmentation (28). The built-in ensembling of nnUNet was utilized for training and inference. Early stopping was implemented such that once the model does not improve on the validation set after 50 epochs, the training is stopped, up to a maximum of 1000 epochs. All other training parameters (learning rate, batch size, data augmentations, loss function) are the default nnUNet settings (28). All training settings described below are trained in the same manner.

#### Adult brain tumor model performance [BraTS model]

Competitions such as Brain Tumor Segmentation (BraTS) Challenge 2012-2021 have shown that deep learning-based solutions can effectively segment adult gliomas, mainly of the high-grade variety (18). Given the proven performance of nnUNet on adult tumors, we sought to determine the performance of these models when applied to the pediatric setting. We hypothesized that, given significant biological and morphological differences between adult and pediatric gliomas, adult tumor-trained model performance would degrade in the pediatric setting. To test this hypothesis, an nnUNet model was trained using the BraTS 2021 training dataset, which contains 1,251 MRI scans and associated expert annotations. To compare performance in the adult and pediatric settings, inference was performed first on an adult hold-out set from BraTS 2021. Inference was then completed on the internal CBTN pediatric test set (n=60).

#### Training from scratch [Scratch model]

Given the similarities in data distribution, we hypothesized that a model trained on pediatric brain tumors would perform better in a pediatric test set than a model trained on adult tumors. To test this hypothesis, using the CBTN training dataset (n=148), an nnUNet model was trained in the same way as the adult model. This model employed only the limited pediatric data available and, as such, relied on much less training data.

#### In-domain transfer learning from adults [Transfer model]

Transfer learning involves leveraging data from other sources, that is often out of distribution, domain, or modality, to initialize models with foundational knowledge that can be then fine-tuned to a particular task (21). This technique has been employed successfully in several medical imaging applications and is commonly performed by leveraging models trained on the 2D ImageNet (29). More recently, 3D medical imaging-specific pretrained models have become available and have demonstrated improved performance compared to ImageNet-trained models on medical tasks. Multiple studies suggest that the closer the data distribution between pretrained neural network and the task, the better (30). Thus, in the case of limited data scenario of pLGG segmentation, we sought to leverage in-domain transfer learning from the adult model combined with pediatric data. Specifically, the nnUNet model is initialized (i.e., pretrained) with the *Adult* model weights and then fine-tuned with additional training on the CBTN data under the same procedure as the *Scratch* model.

#### Stepwise Transfer Learning

Due to the limited quantity of pediatric data available for training, we hypothesized that reducing the number of parameters a network must optimize may help improve model convergence. This can be achieved by freezing specific model parameters during the fine-tuning process. This effectively reduces the complexity of the network for the optimizer. When the model is optimizing both the encoder and decoder simultaneously, it may not be able to find the optimal solution given the sparsity of data. Starting from the checkpoint of *Transfer model*, training is continued with all parameters frozen in either the nnUnet encoder block (*Transfer-Encoder* model) or decoder block (*Transfer-Decoder* model).

### Model evaluation and statistical analysis

The primary performance endpoint of the study was the Dice similarity coefficient (DSC) (31), which measures the relative overlap of the predicted and ground-truth segmentations, with a DSC of >0.80 considered to be worthy of clinical testing (10). DSC was calculated based on Equation [1]:

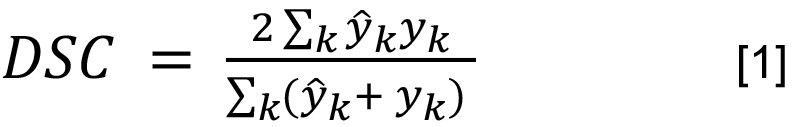

where y_k_ is the ground truth for voxel k of one image, and *ŷ*_*k*_ is the prediction for voxel k of that image. Median DSCs between models were compared with Wilcoxon Rank Sum tests within the CBTN test set. The highest performing model on the CBTN test set was then externally validated on the BCH dataset (n=60). Additionally, 3D volumetric measurement was calculated from each predicted and ground-truth segmentation, and median relative volume difference (RVD) and absolute volume difference (AVD) were evaluated. Other secondary endpoints included aggregated DSC, which calculates DSC over the entire test set population (rather than case-by-case), and intraclass coefficient (ICC) for calculated volumes. Aggregated DSC (DSC_agg_) was calculated based on Equation [2]:

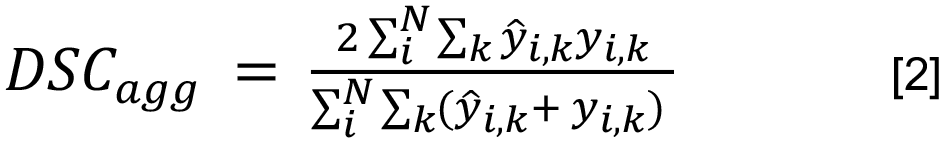

where N is the number of test images, y_i,k_ is the ground truth for voxel k and image i, and *ŷ*_*i*,*j*_ is the prediction for voxel k and image i. Means were compared with ANOVA. All tests were two-sided with a *p*<0.05 considered statistically significant. Statistical analyses were conducted using the Scikit-learn and SciPy packages in Python v3.8. Three inter-rater group DSCs were combined between three experts, between model 1 and experts, and between model 2 and experts. Mean inter-rater group DSCs between combined expert models and two AI models were compared with One-way ANOVA. Mean ratings of three individual experts, combined experts, and two individual segmentation models were compared by One-way ANOVA. Tukey’s ‘Honest Significant Difference’ method was used as post-hoc analysis to find specific groups with significantly different mean DSCs or mean ratings from other groups. All tests were two-sided with a p<0.05 considered statistically significant. Statistical analyses were conducted using R.

### Randomized, blinded clinical acceptability testing and inter-expert variability

While DSC is an important quantitative measure of segmentation performance, positioning the algorithm for real-world use requires clinical validation and benchmarking (32,33). To determine segmentation variation across expert clinicians and diagnosticians of different specializations, a second radiation oncologist specializing in central nervous system tumors and a pediatric neuroradiologist were enlisted to annotate the entire BCH external dataset (n=100) for clinical acceptability testing (Fig. 1B). Pairwise inter-expert variability between the three annotators as well as two deep learning models (*BraTS* model and *Transfer-Encoder* model) was similarly evaluated with DSC to determine if model performance was within the range of inter-expert variability. To assess the clinical utility of the AI models, the three experts conducted a blinded, segmentation rating and acceptability study (Fig. 1C). For each of the 100 BCH cases, each expert was presented with three different segmentations overlaid with each other, with the ability to hide/unhide each individual segmentation as needed (Supplementary Protocol). The three segmentations consisted of at least one, and up to two expert segmentations (from the other two annotators) and at least one, and up to two AI-generated generated segmentations (selected from the *BraTS model* and/or the *Transfer-Encoder* model), selected at random (n=300 total segmentations per reviewer). The expert reviewers were blinded to the origin of the segmentations. The order and color of the segmentations displayed for each scan was randomized to reduce bias. The review was carried out using 3D Slicer (www.slicer.org). Experts were given written instructions and asked to rate each of the three segmentations on a scale from 1 (worst) to 10 (perfect), with a 7 being defined as a clinically acceptable segmentation for volumetric assessment (Fig. S6). For each segmentation, raters were also asked to choose whether the segmentation was AI-generated, also known as the Turing test.

**Figure 1.**
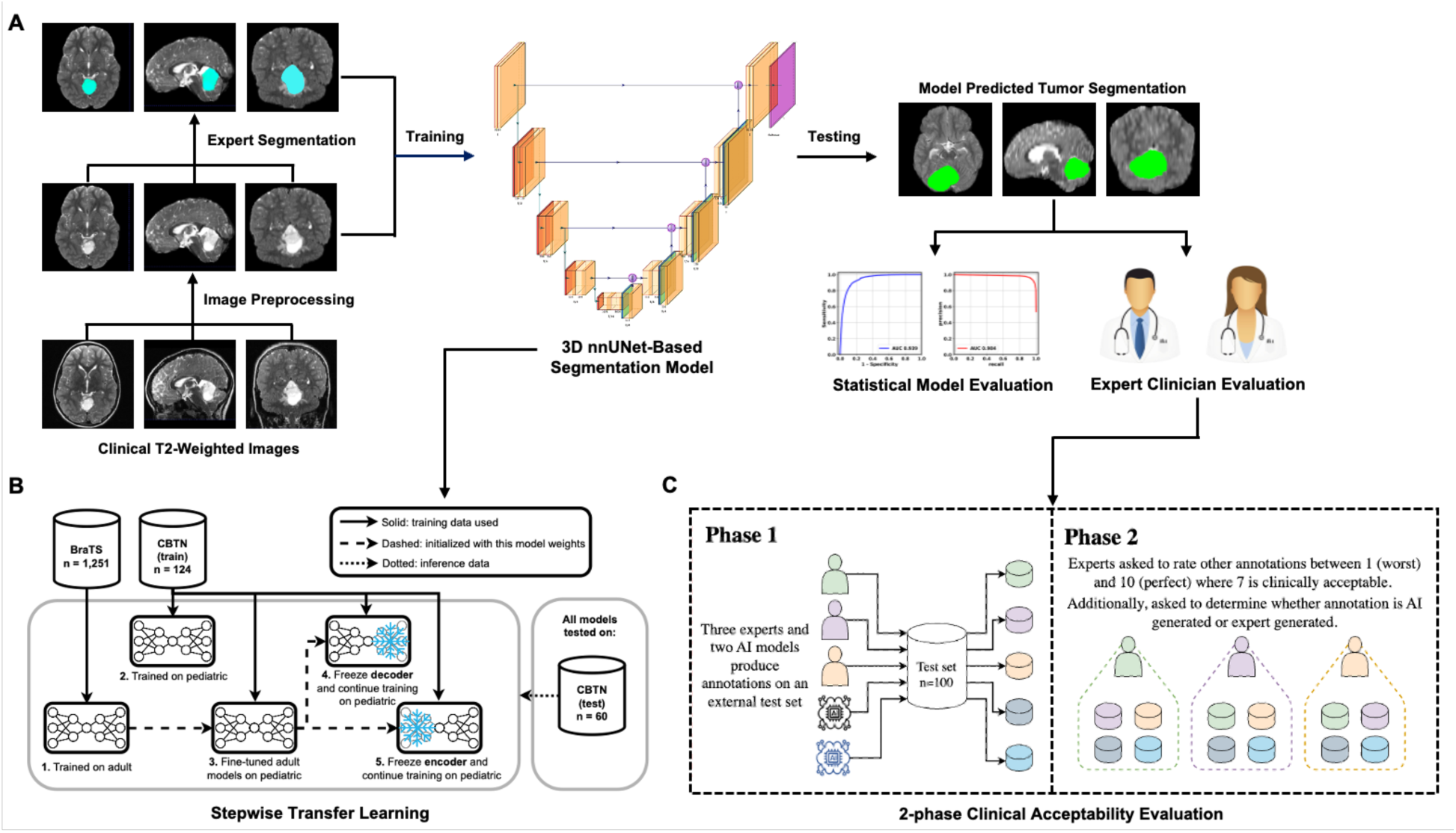
Schematic illustration of the study design. (A) An overview of the study workflow, including data preprocessing, expert segmentation on the tumors, model training/testing and well as the model clinical acceptability evolution. (B) A workflow showing the proposed in-domain stepwise transfer learning with detailed sequential steps involved in this approach. (C) A workflow detailing the 2-phase clinical acceptability evaluation.

## RESULTS

### Patient Characteristics

The total pLGG patient cohort consisted of 284 pLGG patients from two cohorts, with 184 patients in the development set from CBTN cohort and 100 patients in the external test set from BCH (Table 1). Median age was 7 (range: 1-23) in the CBTN cohort and 8 (range: 1-19) in the BCH cohort. 84 patients (45.7%) were female in CBTN cohort, while 53 patients (53%) were female in BCH cohort. All patients had pathologically diagnosed grade I/II low-grade glioma, with a mixture of histologic subtypes of pilocytic astrocytoma (32.4%), optic pathway glioma (10.6%), juvenile pilocytic astrocytoma (4.6%), fibrillary astrocytoma (4.6%), ganglioglioma (4.2%), diffuse astrocytoma (4.2%), and other low-grade glioma/astrocytoma (44.0%). The primary tumor locations were posterior fossa (25.7%), temporal lobe (10.9%), Suprasellar (9.5%), frontal lobe (3.2%), and others (50.5%).

**Table 1.** Patient demographics.

|  | Patient Cohorts |  |
| --- | --- | --- |
|  | CBTN (n = 184) | BCH (n = 100) |
| **Age (years)** |  |  |
| median (range) | 7 (1 - 23) | 8 (1 – 19) |
| **Sex n (%)** |  |  |
| Female | 84 (45.7%) | 53 (53%) |
| Male | 94 (51.1%) | 47 (47%) |
| Unknown | 6 (3.2%) | 0 |
| **Race/Ethnicity n (%)** |  |  |
| Non-Hispanic Caucasian/white | 124 (67.4%) | 68 (68%) |
| African American/Black | 24 (13.0%) | 5 (5%) |
| Hispanic/Latinx | 17 | 4 (4%) |
| Asian American/Asian | 3 (1.6%) | 4 (4%) |
| Other/Unknown | 33 (17.9%) | 19 (19%) |
| **Histologic diagnosis n (%)** |  |  |
| pilocytic astrocytoma | 58 (31.5%) | 34 (34%) |
| Pilomyxoid Astrocytoma | 10 (5.4%) | 0 |
| Juvenile Pilocytic Astrocytoma | 0 | 13 (13%) |
| Ganglioglioma | 1 (0.5%) | 11 (11%) |
| Oligodendroglioma | 1 |  |
| Diffuse astrocytoma | 9 (4.9%) | 3 (3%) |
| Fibrillary Astrocytoma | 13 (7.1%) | 0 |
| Optic pathway glioma | 27 (14.7%) | 3 (3%) |
| Other Low-grade glioma/astrocytoma | 65 (35.3%) | 36 (36%) |
| **Primary Tumor Location n (%)** |  |  |
| Posterior fossa | 45 (24.5%) | 28 (28%) |
| Temporal lobe | 13 (7.1%) | 18 (18%) |
| Frontal Lobe | 7 (3.8%) | 2 (2%) |
| Cerebellum | 0 | 18 (18%) |
| Suprasellar | 21 (11.4%) | 6 (6%) |
| Optic Pathway | 27 (14.7%) | 3 (3%) |
| Brainstem | 9 (4.9%) | 3 (3%) |
| Thalamus | 5 (2.7%) | 2 (2%) |
| Others | 47 (25.5%) | 20 (20%) |

CBTN = Children Brain Tumor Consortium; BCH = Boston Children Hospital.

### In-domain Stepwise Transfer learning with fine-tuning improves performance

The performance of the five model settings outlined in Fig. 1 is illustrated in Table 2. The *BraTS* model performed highly on held-out adult data, however, the performance of the model decreased significantly when applied to the pediatric data (Table 2; median DSC 0.926 [IQR 0.886-0.953] to 0.812 [IQR 0.559-0.888], *p*<0.05). Additionally, accuracy of volumetric assessment declined significantly when the *BraTS* model was applied to pediatric data (Table 2; median RVD: 0.052 [IQR 0.024-0.119] to 0.192 [IQR 0.109-0.682], *p*<0.05). This result highlighted the difference between adult gliomas and pediatric low-grade gliomas and emphasized the importance of developing a model capable of accurately segmenting pediatric cases. The *Scratch* model and all *Transfer* models performed significantly better (*p*<0.05 for each) than the *BraTS* model, with median DSC of 0.862 [IQR 0.672-0.91], 0.871 [IQR 0.724-0.914], 0,877 [IQR 0.708-0.914] and 0.877 [IQR 0.715-0.914] for *Scratch, Transfer, Transfer-Decoder*, and *Transfer-Encoder* models, respectively (Table 2 & Fig. 2). Of all approaches investigated, the highest performing was the *Transfer-Encoder* model (Table 2; median DSC: 0.877 [IQR 0.715-0.914]; median relative volume difference (RVD): 0.109 [IQR 0.032-0.31]). While both fine-tuned transfer learning models showed near equivalent performance for the primary endpoint, we chose *Transfer-Encoder* model for further testing giving its increased aggregated DSC (Table 2; aggregated DSC: 0.730 to 0.840), which indicated it had fewer incidences of empty segmentation masks. Overall, *Transfer-Encoder* model has the lowest empty mask percentage.

**Table 2.** Model performance for internal test set (n=60).

| Models | Median DSC (IQR) | Aggregated DSC | Median RVD (IQR) | ICC | Percentage of Cases with DSC 0 |
| --- | --- | --- | --- | --- | --- |
| *BraTS* model | 0.812 (0.559-0.888) | 0.730 | 0.192 (0.109-0.682) | 0.736 | 16.7% |
| *Scratch* model | 0.862 (0.672-0.91) | 0.815 | 0.098 (0.04-0.407) | 0.804 | 11.7% |
| *Transfer* model | 0.871 (0.724-0.914) | 0.823 | 0.124 (0.036-0.334) | 0.799 | 10% |
| *Transfer-Decoder* model | 0.877 (0.708-0.914) | 0.832 | 0.126 (0.052-0.289) | 0.832 | 8.3% |
| *Transfer-Encoder* model | 0.877 (0.715-0.914) | 0.840 | 0.109 (0.032-0.31) | 0.825 | 6.7% |

IQR = inter-quartile; DSC = Dice similarity coefficient; RVD: relative volume difference; ICC = intra-class correlation coefficient.

**Figure 2.**
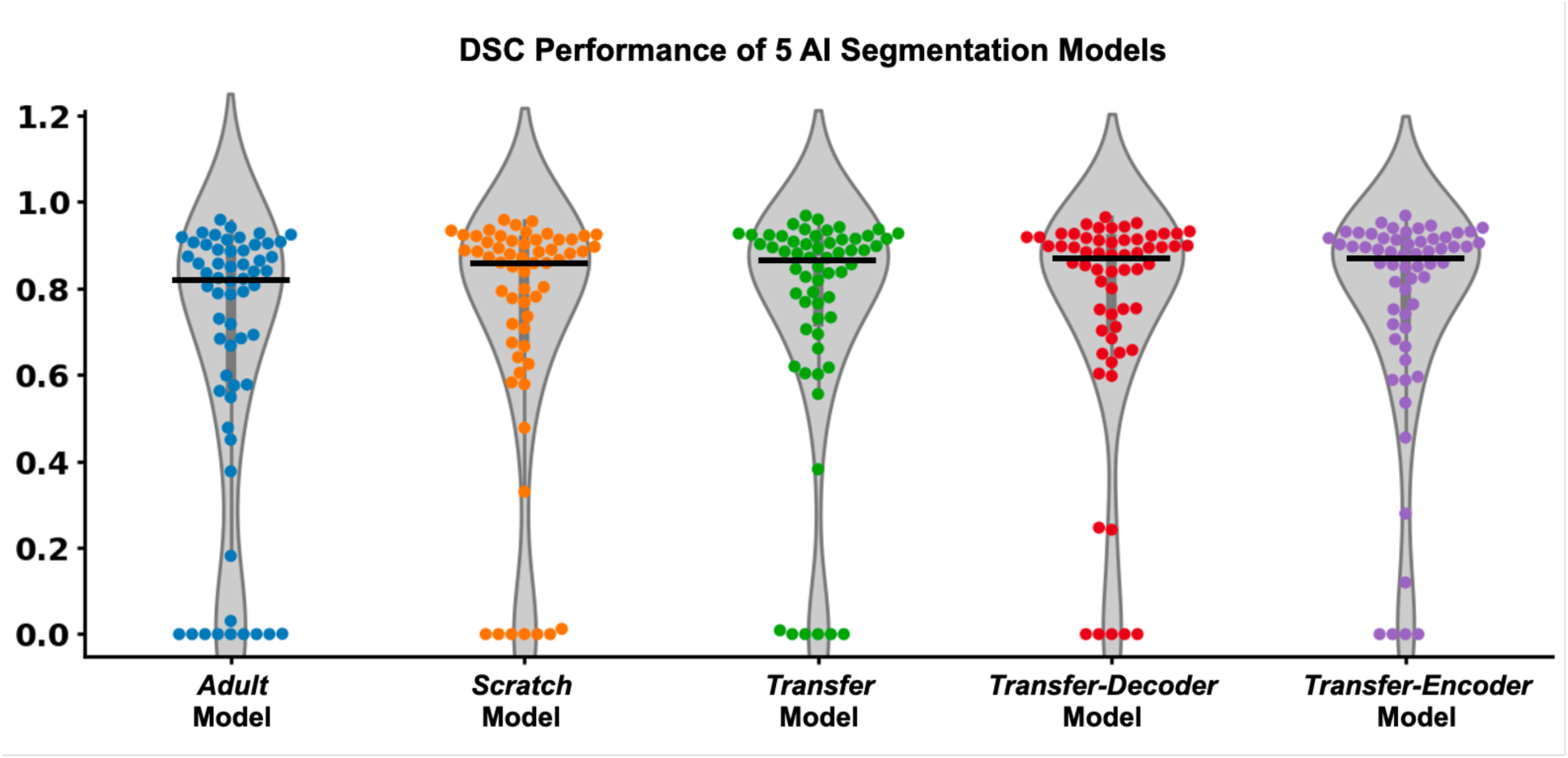
Comparative performance of deep learning training methodologies on internal validation set (n=60). Among the five different segmentation models assessed, the methods using stepwise transfer learning (Transfer-Decoder and Transfer-Encoder) had the highest segmentation accuracy. Additionally, the Transfer-Encoder model generated the fewest segmentations with a DSC: 0 (n=4, 6.7%) (indicating a complete segmentation miss). Conversely, the BraTS model exhibited the highest number of segmentations with a DSC: 0 (n=11, 18.3%). The Transfer-Encoder model demonstrated the highest median DSC (0.877 [IQR 0.715-0.914]), and was selected for further investigation. The BraTS model, trained only on adult glioma, demonstrated the lowest median DSC (0.812 [IQR 0.559-0.888]).

### External validation of Stepwise Transfer Learning

On an external testing set with expert segmentations (n=60), the *Transfer-Encoder* model achieved median DSC 0.833 [IQR 0.743-0.900] and median RVD of 0.161 [IQR 0.058-0.393] as compared to manual segmentations. We performed failure analysis for cases with DSC<0.6 and found 6 cases in total. The failures were caused by: 1) tumor located in ventricle (Fig. 3E; n=1); 2) large cystic area in brain (Fig. 3A, C&D; n=3); empty segmentation from poor image quality due to respacing for large slice thickness (Fig. 3B; n=1); 4) under-segmentations for large heterogeneous tumor lesion (Fig. 3F; n=1).

**Figure 3.**
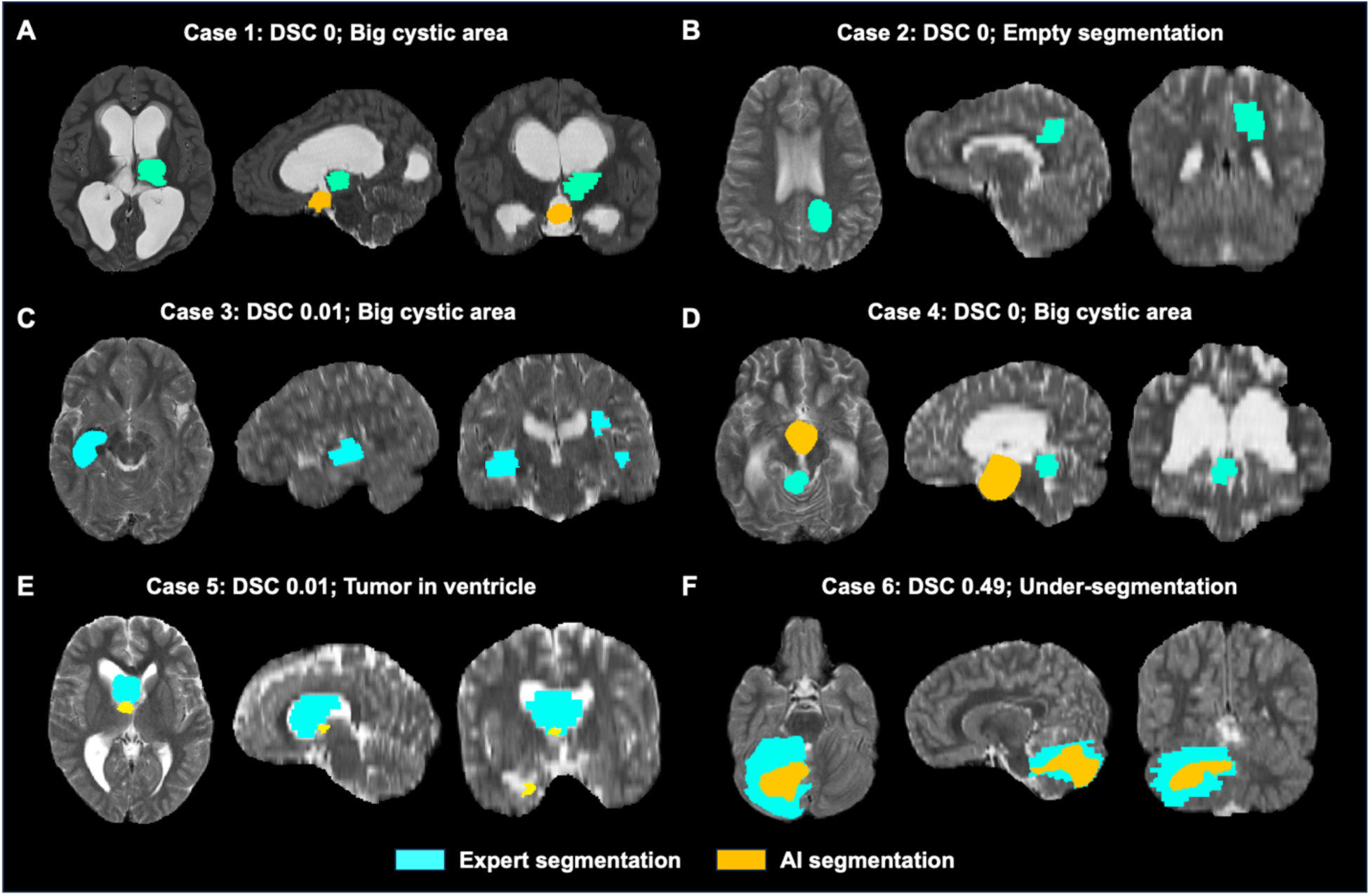
Analysis of Transfer-Encoder model failures. Failure analysis was performed based on cases with DSC<0.6 and 6 cases were identified in total. The failures were caused by big cystic area in brain (A, C&D), empty segmentation from poor image quality due to respacing for large slice thickness (B), tumor located in ventricle (E), under-segmentations for large heterogeneous tumor lesion (F).

### Stepwise transfer learning demonstrates clinical expert-level performance

The distribution of inter-rater DSCs between experts and AI models is presented in Fig. 4A, while the heatmap in Fig. 4B displays the median inter-rater DSCs for each pair. We further compared the inter-rater DSCs from three experts, the *Transfer-Encoder* model, and the *BraTS* model. The *Transfer-Encoder* model (median: 0.834 [IQR 0.726-0.901]) did not show a significant difference (p=0.13) compared to the three experts (median: 0.861 [IQR 0.795-0.905]), but it exhibited a significantly higher DSC (p<0.01) than the *BraTS* model (median: 0.790 [IQR 0.662-0.870]) (Fig. 4C).

**Figure 4.**
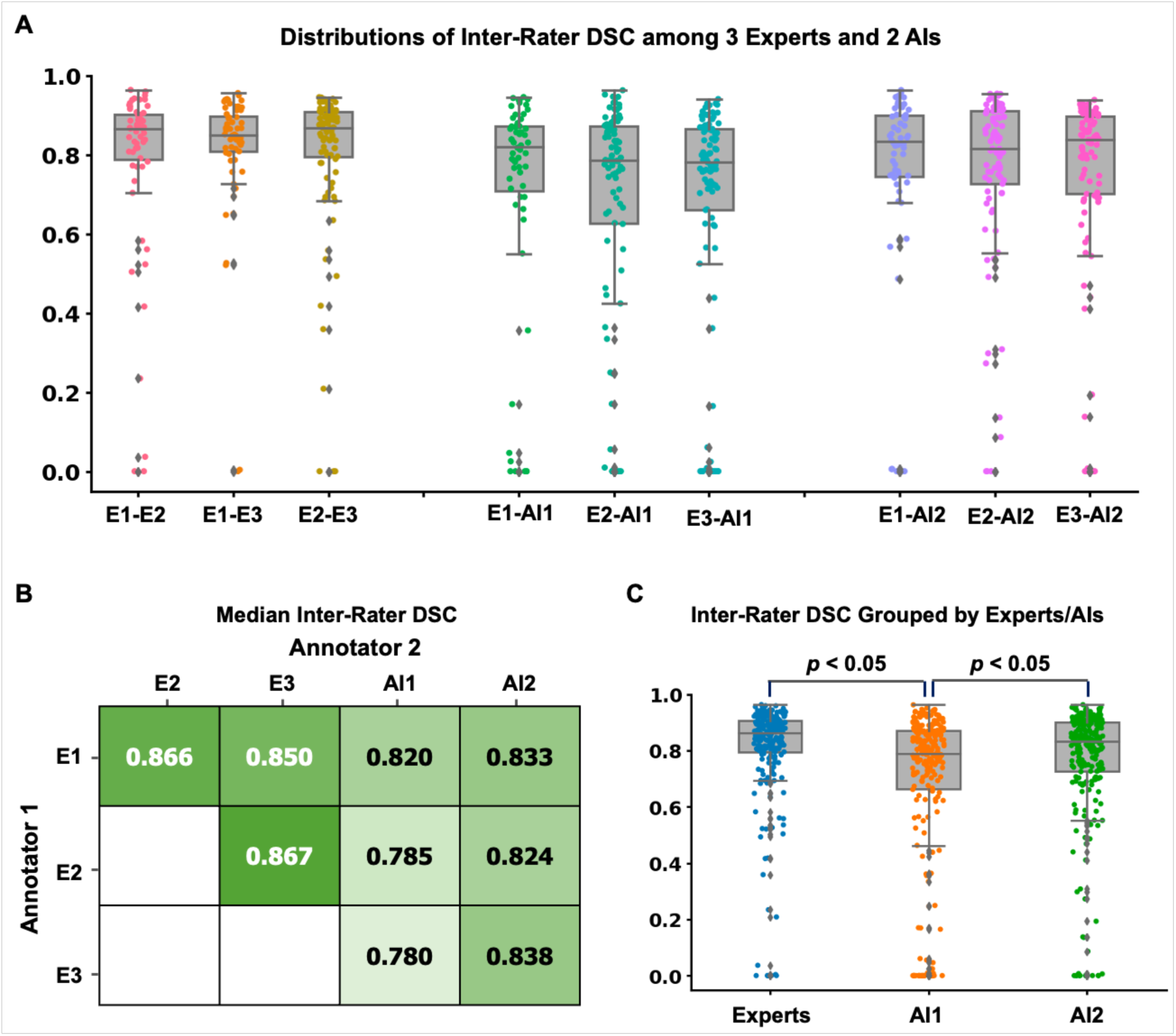
Clinical acceptability testing. Three human experts were invited to rate randomized AI-generated (BraTS model or Transfer-Encoder model) or expert-generated segmentations blinded to segmentation origin (n=100 scans, 300 segmentations). (A) The distributions of inter-rater DSCs from three experts and two AI models for external test dataset (n=100). (B) The median inter-rater DSCs between each pair of the experts and AI models. (C) Boxplot shows inter-rater DSCs grouped by experts, Transfer-Encoder model and BraTS model. Both Transfer-Encoder model and average experts shows significantly higher (p<0.05) inter-rater DSCs than BraTS model. There was no significant difference on inter-rater DSCs between average experts and Transfer-Encoder model. DSC: Dice similarity coefficient; AI: artificial intelligence; E1: Expert 1; E2: Expert 2; E3: Expert 3; AI1: BraTS model; AI2: Transfer-Encoder model.

For clinical acceptability testing, the segmentation ratings of the *Transfer-Encoder* model (median: 9 [IQR 7-9]) were significantly higher (*p*<0.01 for each) than those of Expert 1 (median: 7 [IQR 6-9]), Expert 3 (median: 7 [IQR 5-8]), the average expert (median: 7 [IQR 6-9]), and the *BraTS* model (median: 8 [IQR 6-9]). However, there was no significant difference between the *Transfer-Encoder* model and Expert 2 (median: 8 [IQR 7-9]) (Fig. 5A). Expert 2 had significantly higher ratings (*p*<0.01 for each) compared to Expert 1, Expert 3, and the average experts. There was no significant difference (p=0.54) between Expert 1 and Expert 3 in terms of the ratings (Fig. 5A). Furthermore, the Transfer-Encoder model demonstrated a significantly higher (*p*<0.05 for each) proportion of clinically acceptable segmentations (rating score≥7) compared to two out of three experts (Expert 1 & Expert 3) and the experts average (Fig. 5B: *Transfer-Encoder*: 80.2%; *BraTS*: 72.1%; Expert 1: 68.3%; Expert 2: 78.7%; Expert 3: 49.3%; Experts average: 64.8%). Finally, results from the Turing test revealed consistently low accuracy in distinguishing AI-generated segmentations from those produced by experts. The Transfer-Encoder model segmentations were correctly identified by experts in only 26.0% of scans, which is lower than the correct identification rates for Expert 1 (35.0%), Expert 2 (66.5%), Expert 3 (26.9%), and the *BraTS* model (41.5%) (Fig. 5C).

**Figure 5.**
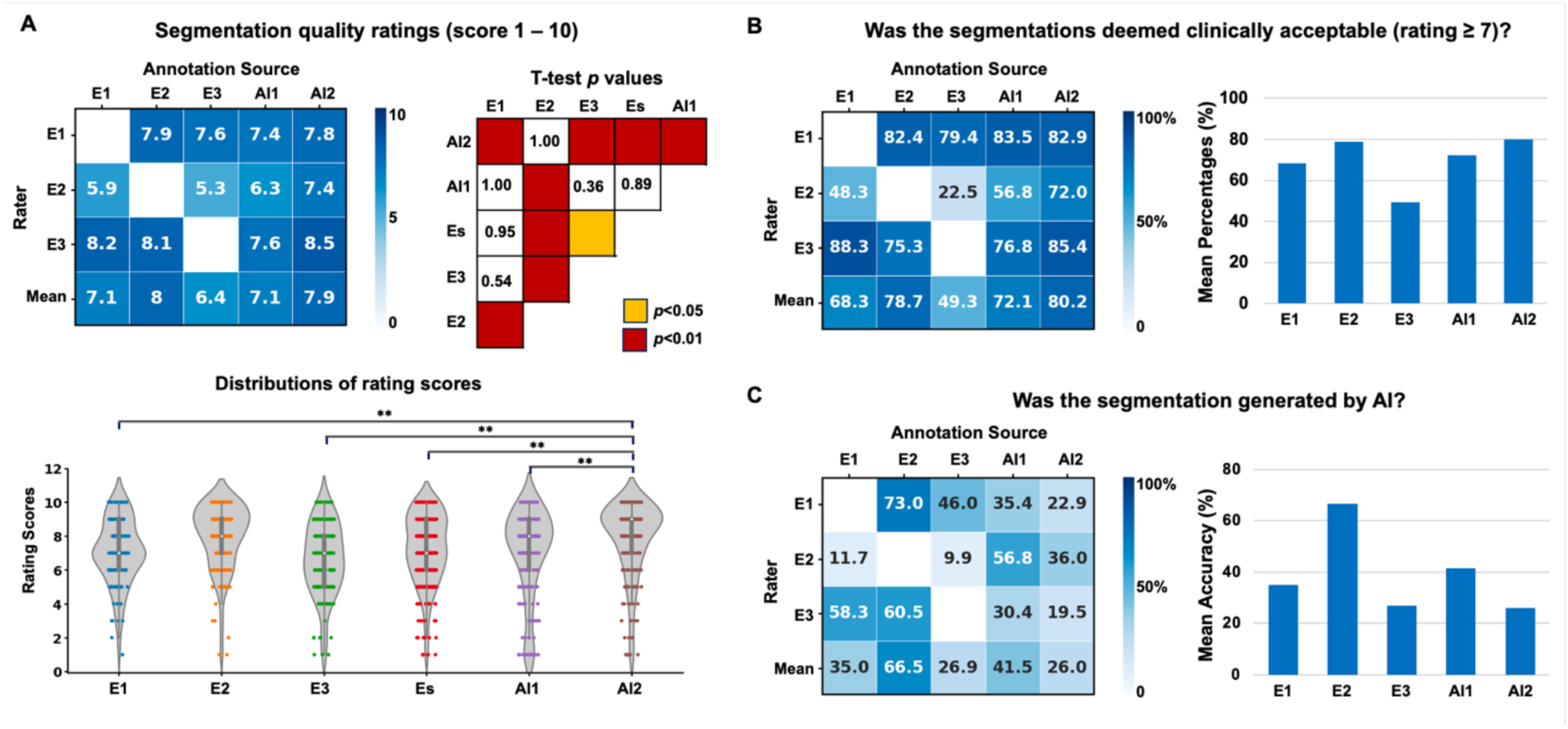
(A) The mean segmentation rating scores for the three experts and two AI models, as evaluated by the experts, are presented in a heatmap. Violin plots are used to compare and group the rating scores for each expert and AI model, along with the corresponding p-values from statistical tests for group comparisons. (B) The percentage of segmentations that were considered acceptable, with a rating score of 7 or above, is summarized for each expert and model. (C) The accuracy of determining whether annotations were AI generated is shown for each expert and model, indicating how well the experts were able to distinguish between AI-generated and expert-generated segmentations. DSC: Dice similarity coefficient; AI: artificial intelligence; E1: Expert 1; E2: Expert 2; E3: Expert 3; AI1: BraTS model; AI2: Transfer-Encoder model. DSC: Dice similarity coefficient; AI: artificial intelligence; E1: Expert 1; E2: Expert 2; E3: Expert 3; Es: average experts; AI1: BraTS model; AI2: Transfer-Encoder model.

## DISCUSSION

In this study, we developed, validated, and clinically benchmarked a deep learning pipeline using stepwise transfer learning for automated, expert-level pLGG segmentation and volumetric measurement. Accurate tumor auto-segmentation models could be useful for risk-stratification, monitoring tumor progression, assessing treatment response, and surgical approach (6), though have had limited traction in pediatric tumors due to very sparse available training data. We leveraged a novel strategy of in-domain, stepwise transfer learning to demonstrate measurable gains in segmentation accuracy and clinical acceptability that was on par with clinician performance. To our knowledge, this is the first study to utilize stepwise transfer learning in this context and to evaluate clinical acceptability of auto-segmentation tools. The rigorous clinical benchmarking studies with three blinded experts suggest that this approach is nearing a performance ceiling for pLGG segmentation – i.e., output is comparable and indistinguishable to human experts. These findings position the model for prospective testing and clinical translation.

The current state-of-the-art approaches for automated brain tumor segmentation rely on deep learning. However, most available auto-segmentation tools have been specifically developed and trained for adult brain cancers, particularly glioblastoma (GBM) (9,18,19). In this work, we find that tools such as these do not effectively generalize to pediatric brain tumors. Performance degradation may stem from the distinctive heterogeneous imaging appearance and types of pediatric brain tumors compared to adult brain tumors, as well as the anatomical differences resulting from the ongoing brain development in children. Several studies have proposed various DL solutions to address the segmentation of pediatric brain tumors, achieving Dice similarity coefficients (DSC) ranging between 0.68 and 0.88 (6,16,34,35), however the clinical acceptability of these approaches was not validated. To date, only one study has proposed an algorithm for pLGG segmentation, achieving a DSC of 0.77 (17). This study utilized FLAIR images from 311 patients from a single institution. The proposed model employed deep Multitask Learning, incorporating a tumor’s genetic alteration classifier as an auxiliary task to the main segmentation network (17). However, this model was trained on a limited number of MRI scans from a single institute and lacked external validation and clinical testing. In the present study, our stepwise transfer learning model achieved median DSC of 0.833 [IQR 0.743-0.900] with a median RVD of 0.161 [IQR 0.058-0.393] in the external test set, which represents a significant improvement over previous work (17). Improved performance may be due to sequential knowledge transfer - first from the adult setting, and then the pediatric setting. Additionally, freezing the encoder or decoder in the final fine-tuning step enabled optimization of a smaller parameter space, which may have mitigated overfitting given the limited amount of data. Training on a sufficient quantity of pediatric data from scratch may obviate the need for transfer learning, but what represents “sufficient” is yet to be defined for pLGGs, and current datasets remain relatively small.

While statistical metrics like the DSC and RVD offer valuable insights into a model’s overall segmentation performance, it is important to acknowledge their limitations in providing a comprehensive evaluation of a model’s utility (6). To ensure a thorough evaluation and facilitate the clinical translation of our model, we conducted a rigorous clinical-acceptability evaluation and validation process involving three expert clinicians. This multidimensional evaluation approach captures additional nuances and considerations that impact the model’s practical utility in a real-world clinical setting. The involvement of expert clinicians provides valuable feedback and insights, accelerating the translation of the model into clinical practice. In our study, we went a step further by conducting blinded, segmentation rating, acceptability, and Turing tests involving three expert clinicians. Notably, all experts performed worse than random chance (50%) in predicting the origin of the transfer learned model segmentations, suggesting that this model passes the Turing test, though this was not the case for the adult-trained model. To our knowledge, this is the first brain tumor segmentation study to incorporate such a clinical-acceptability test. The results highlight the importance of such clinical testing in positioning a model for potential clinical translation.

There are several limitations of this study that should be taken into consideration. Firstly, the study is retrospective in nature and selection of scans for inclusion for this study, while performed a priori and based solely on availability, may introduce bias. Secondly, the model utilizes only T2W images, as these were the most commonly available for all the patients included in our analysis. Although T2 FLAIR images are commonly used for defining tumor regions, T2 FLAIR images were not available for many patients with pLGGs in our study, particularly in the CBTN cohort. Consequently, it becomes challenging to distinguish vasogenic edema from the tumor region on T2W images, making it difficult to accurately segment tumor areas based solely on T2W images. Despite these challenges, our study demonstrated that our DL model exhibited impressive segmentation performance compared to expert clinicians, suggesting that the model successfully captured the feature differences. However, we acknowledge that the incorporation of multiple MRI modalities, such as T2W, pre-contrast T1W, post-contrast T1W, and T2 FLAIR images, would enhance the granularity of tumor segmentation in pLGG patients. On the other hand, an advantage of a T2W-only model, is that it may be more widely applicable in situations where multiparametric and contrast-enhanced scans are unavailable, which can sometimes be the case for low-grade glioma studies. Furthermore, it is important to note that our study focused solely on whole tumor segmentation, and not the segmentation of specific tumor subregions. Consequently, the clinical utility of our findings may be restricted in certain cases, when change in cystic component is not relevant to the clinical response assessment. Finally, it is notable that the algorithm did fail on some cases, and while we identified certain factors that were associated with failures, it is difficult to predict with certainty why a failure occurred owing to the black-box nature of deep learning algorithms. Therefore, it is important for the model output to undergo a clinical review prior to use in clinical decision-making.

## CONCLUSION

We developed, externally validated, and clinically benchmarked an automated deep learning pipeline using in-domain, stepwise transfer learning that enables expert-level MRI segmentation of pediatric low-grade gliomas. On blinded evaluation, the model demonstrated clinically acceptable performance that was higher on average than clinical experts. Prospective and longitudinal evaluation of the pipeline is planned to determine the algorithm’s potential for integration into the clinical care of children with low-grade glioma.

## Abbreviations

AUC: area under the curve
CNN: convolutional neural network
DICOM: Digital Imaging and Communications in Medicine
pLGG: pediatric low-grade glioma
DSC: dice similarity coefficient.

## Funding

This study was supported in part by the National Institutes of Health (NIH) (U24CA194354, U01CA190234, U01CA209414, R35CA22052, and K08DE030216), the National Cancer Institute (NCI) Spore grant (2P50CA165962), the European Union – European Research Council (866504), the Radiological Society of North America (RSCH2017), the Pediatric Low-Grade Astrocytoma Program at Pediatric Brain Tumor Foundation, and the William M. Wood Foundation.

## Competing Interests

All the authors declare no competing interests.

## Author Contributions

Study design: A.B., B.H.K.; code design, implementation and execution: A.B.; acquisition, analysis or interpretation of data: A.B., Z.Y., S.V., R.C., B.H.K.; image annotation: B.H.K., S.P., M.T.; writing of the manuscript: A.B., Z.Y., B.H.K.; critical revision of the manuscript for important intellectual content: all authors; statistical analysis: Z.Y., Y.Z.; study supervision: B.H.K., H.J.W.L.A., T.P., D.H.K.

## Data availability

BraTS data including raw MRI images may be requested from The Cancer Image Archive (https://www.cancerimagingarchive.net). Although raw MRI imaging data cannot be shared, all measured results to replicate the statistical analysis are shared at the GitHub webpage: https://github.com/BoydAidan/nnUNet_pLGG. Furthermore, we include test samples from a publicly available data set with deep learning and expert reader annotations.

## Code availability

The code of the deep learning system, as well as the trained model and statistical analysis are publicly available at the GitHub webpage: https://github.com/BoydAidan/nnUNet_pLGG.

## Notes

### Competing Interest Statement

The authors have declared no competing interest.

### Author Declarations

The Institutional Review Board (IRB) of the Dana-Farber/Boston Children's/Harvard Cancer Center gave ethical approval for this work.

### Summary of Updates

author names updated

