## supplement for "Expert-level pediatric brain tumor segmentation in a limited data scenario with stepwise transfer learning" (from another version of this preprint: version 1, 2023-06-30)

### Supplementary Materials

#### Supplementary Tables

**Table S1.** MR scanner manufacturers and models used for brain scans from Children Brain Tumor Network (CBTN) cohort.

**Table S2.** MR scanner manufacturers and models used for brain scans from Boston Children's Hospital (BCH) cohort.

#### Supplementary Figures

**Figure S1.** Slice thickness of CBTN cohort and BCH cohort.

**Figure S2.** Magnetic fields of MR scanners of CBTN cohort and BCH cohort.

**Figure S3.** Repetition times of T2-weighted images from CBTN cohort and BCH cohort.

**Figure S4.** Echo times of T2-weighted images from CBTN cohort and BCH cohort.

**Figure S5.** Example of review chart for expert clinical validation and acceptability testing for tumor segmentation.

**Figure S6**. Representative examples of manual tumor segmentations from three expert clinicians.

#### Supplementary Protocol: Clinical Validation and Acceptability Testing for Tumor Segmentation

**Supplementary Tables**

**Table S1.** MR scanner manufacturers and models used for brain scans from Children Brain Tumor Network (CBTN) cohort.

| Manufacturer | Model | Patients N (%) |
|---|---|---|
| SIEMENS | Avanto | 2 (1.09%) |
| SIEMENS | Skyra | 26 (14.13%) |
| SIEMENS | Symphony | 4 (2.17%) |
| SIEMENS | TrioTim | 7 (3.80%) |
| SIEMENS | Prisma | 29 (15.76%) |
| Unspecified | Unspecified | 116 (63.04%) |

**Table S2.** MR scanner manufacturers and models used for brain scans from Boston Children's Hospital (BCH) cohort.

| Manufacturer | Model | Patients N (%) |
|---|---|---|
| GE Medical Systems | GENESIS_SIGNA | 16 (16.33%) |
| GE Medical Systems | SIGNA EXCITE | 24 (24.49%) |
| GE Medical Systems | SIGNA HDx | 1 (1.02%) |
| GE Medical Systems | Signa HDxt | 8 (8.16%) |
| Philips Medical Systems | Intera | 1 (1.02%) |
| Philips Medical Systems | Panorama HFO | 1 (1.02%) |
| SIEMENS | Avanto | 1 (1.02%) |
| SIEMENS | Espree | 2 (2.04%) |
| SIEMENS | Skyra | 10 (10.20%) |
| SIEMENS | Symphony | 4 (4.08%) |
| SIEMENS | TrioTim | 9 (9.18%) |
| SIEMENS | Verio | 1 (1.02%) |
| TOSHIBA | Titan | 1 (1.02%) |
| Unspecified | Unspecified | 19 (19.39%) |

**Supplementary Figures**

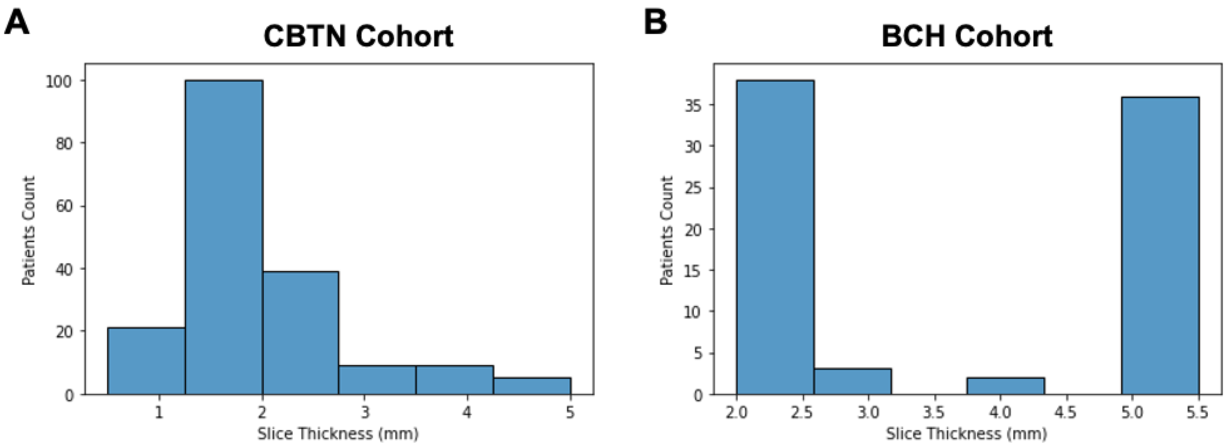


**Figure S1.** Slice thickness of CBTN cohort (A) and BCH cohort (B). CBTN: Children brain tumor network; BCH: Boston Children's Hospital.

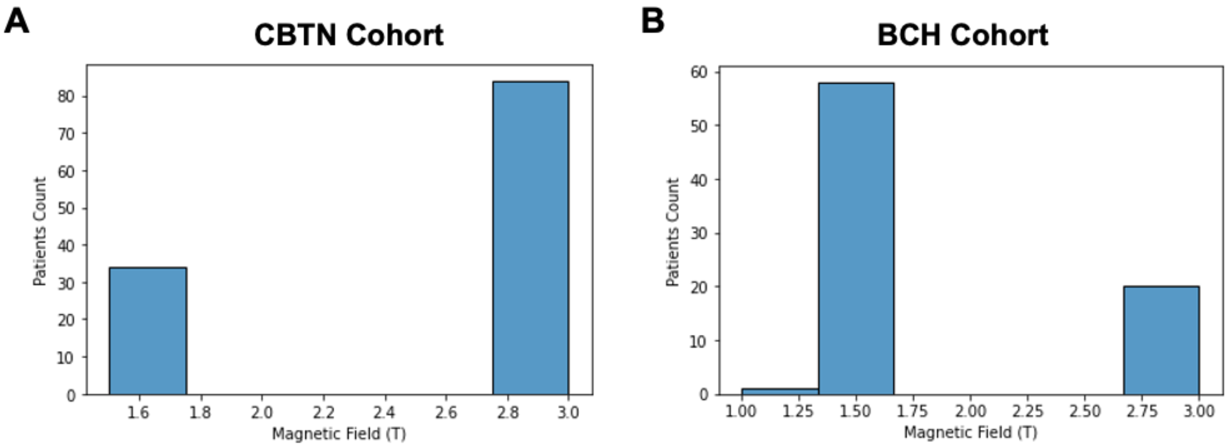


**Figure S2.** Magnetic fields of MR scanners of CBTN cohort (A) and BCH cohort (B). CBTN: Children brain tumor network; BCH: Boston Children's Hospital.

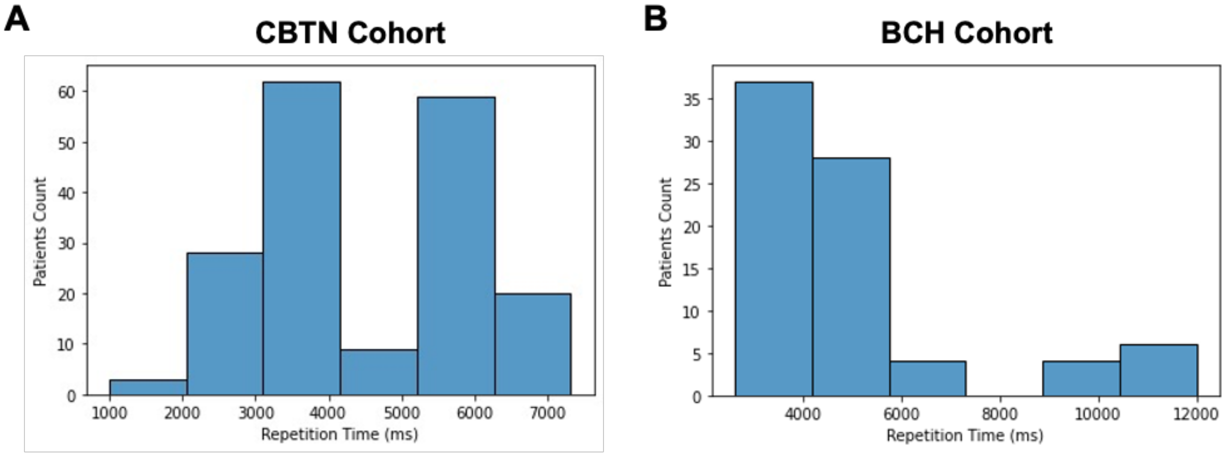


**Figure S3.** Repetition times (TR) of T2-weighted images from CBTN cohort (A) and BCH cohort (B). CBTN: Children brain tumor network; BCH: Boston Children's Hospital.

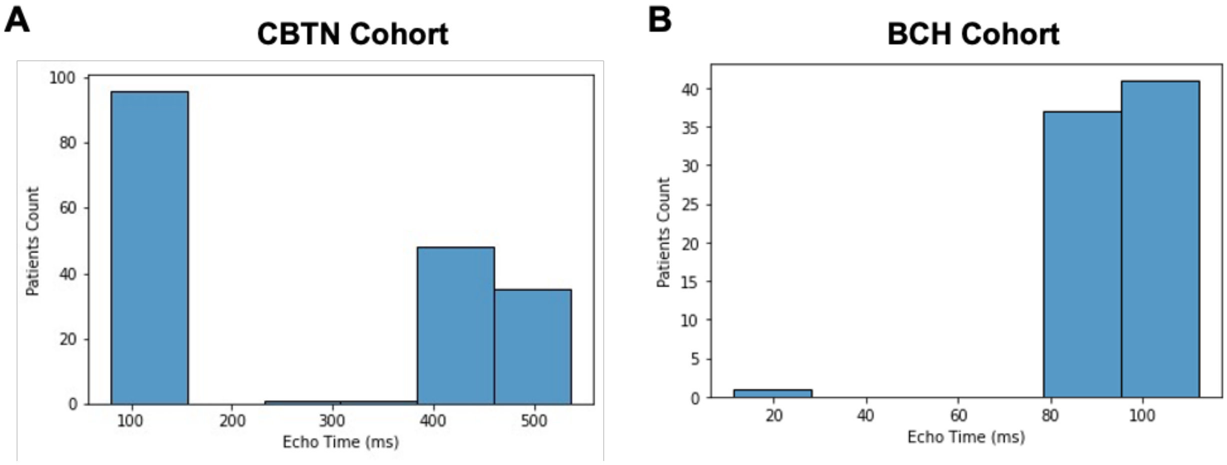


**Figure S4.** Echo times (TE) of T2-weighted images from CBTN cohort (A) and BCH cohort (B). CBTN: Children brain tumor network; BCH: Boston Children's Hospital.

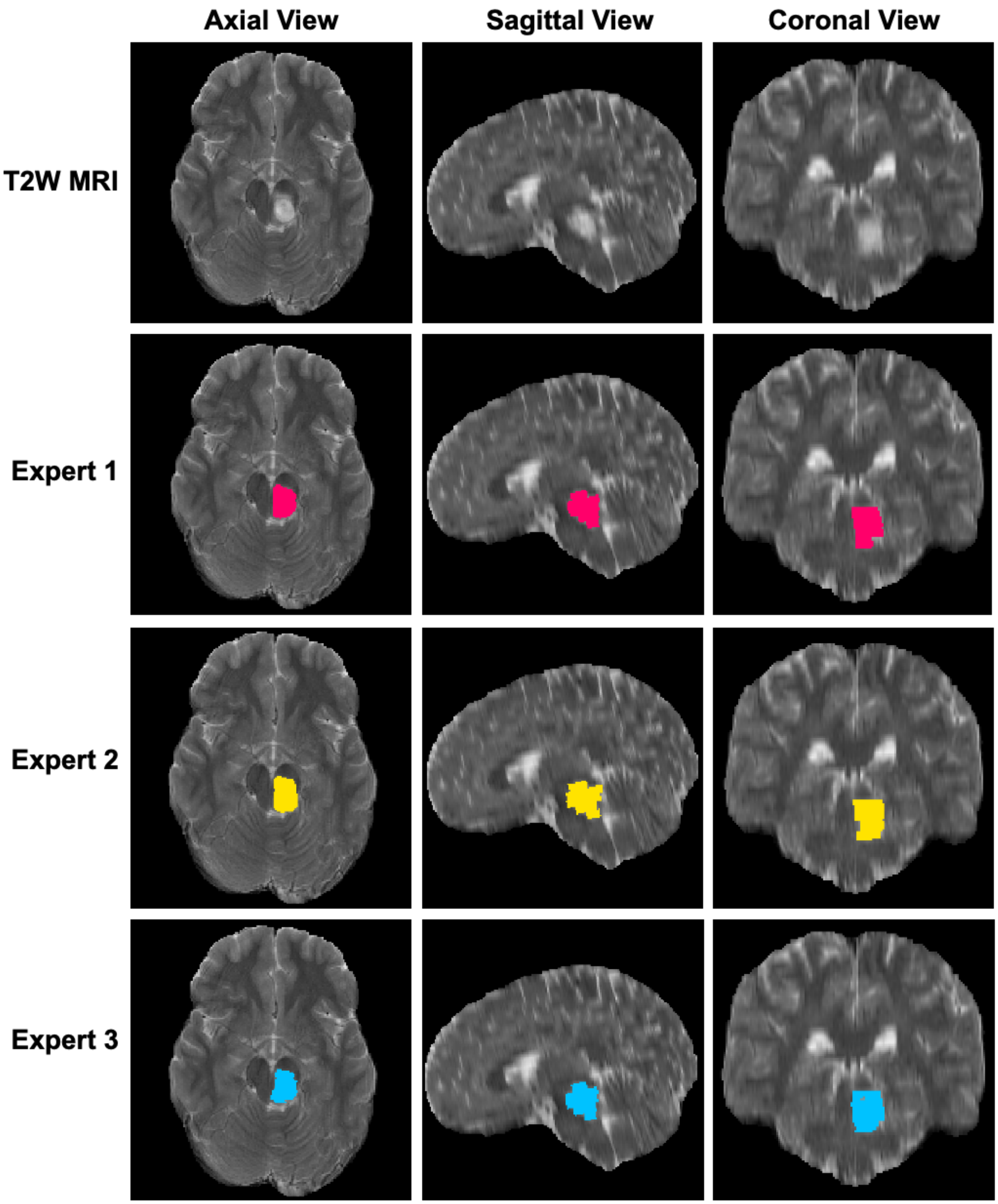


**Figure S5**. Representative examples of manual tumor segmentations from three expert clinicians.

| | | Ranking of segmentations (a,b,c) | | |
|---|---|---|---|---|
| **Scan ID** | | **A** | **B** | **C** |
| 1 | Rating | 7 (acceptable) | 4 | 6 |
| | AI generated? | ☐ | ☑ | ☑ |
| 2 | Usable? | 7 (acceptable) | 1 (worst) | 10 (perfect) |
| | AI generated? | ☑ | ☑ | ☐ |
| 3 | Usable? | 5 | 6 | 5 |
| | AI generated? | ☑ | ☐ | ☑ |
| 4 | Usable? | 9 | 6 | 8 |
| | AI generated? | ☑ | ☑ | ☐ |
| 5 | Usable? | 7 (acceptable) | 5 | 6 |
| | AI generated? | ☐ | ☑ | ☑ |
| 6 | Usable? | 10 (perfect) | 7 (acceptable) | 6 |
| | AI generated? | ☐ | ☑ | ☑ |
| 7 | Usable? | 10 (perfect) | 6 | 9 |
| | AI generated? | ☐ | ☑ | ☑ |
| 8 | Usable? | 1 (worst) | 8 | 7 (acceptable) |
| | AI generated? | ☑ | ☐ | ☑ |
| 9 | Usable? | 10 (perfect) | 7 (acceptable) | 7 (acceptable) |
| | AI generated? | ☐ | ☑ | ☑ |
| 10 | Usable? | 7 (acceptable) | 6 | 8 |
| | AI generated? | ☑ | ☑ | ☐ |

**Figure S6.** Example of review chart for expert clinical validation and acceptability testing for tumor segmentation.

**Supplementary Protocol: Clinical Validation and Acceptability Testing for Pediatric Brain Tumor Segmentation**

Thank you for your participation in this study. This task involves the review, rating, and acceptability designation of human- and AI-generated glioma tumor segmentations on T2-weighted MRI. For each scan, you will be presented with **three** overlaid segmentations, of which **1-2** are AI-generated, and **1-2** are human-generated by a radiation oncologist or neuroradiologist (other than yourself). I have shared the folders with you via DropBox. You will be asked to complete a spreadsheet with the following:

1) Rank each segmentation from 1 to 10 (terrible to perfect). A rating of 7 or higher should be considered "acceptable," meaning "minor modifications needed, but segmentation would be expected to be within 5% of volumetric variation."
2) Designate if you believe the segmentation is AI or human-generated (Turing test).

Please follow the spreadsheet link that will be emailed to you.

Step-by-Step Slicer Instructions as follows:

1) Open Slicer
2) Select "Add Data"

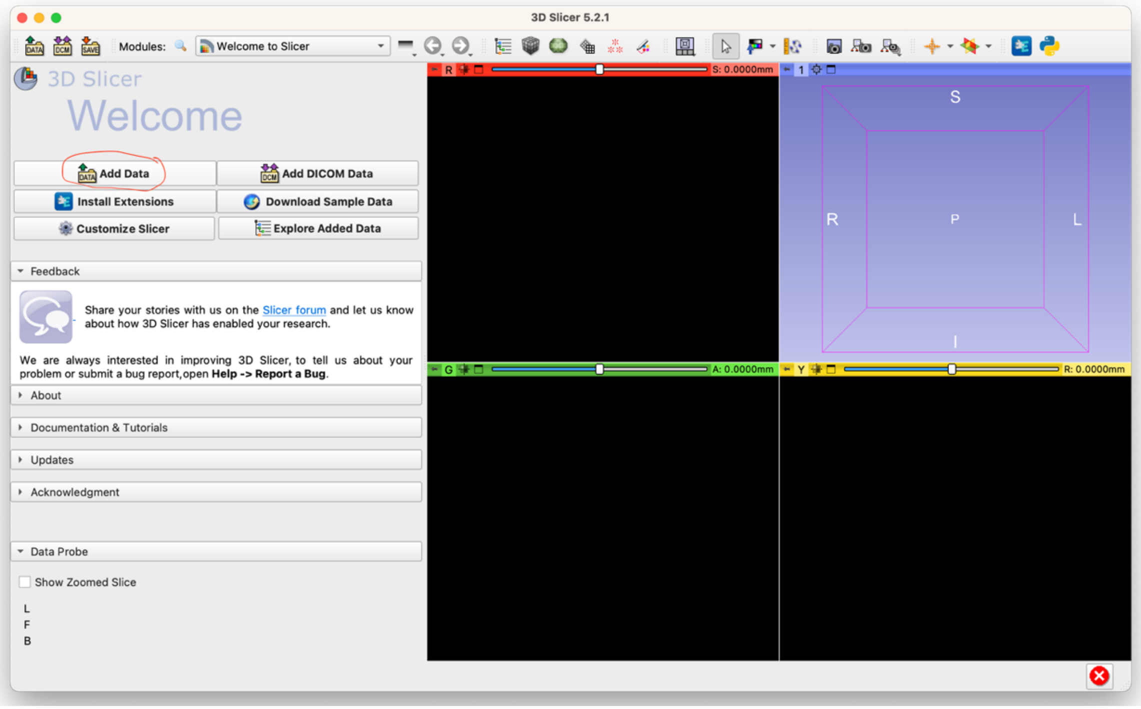

3) Navigate to the Folder containing the scan and segmentations and select “Open.” (Or drag the folder into Slicer and select “Any Data”> OK). You should see this:

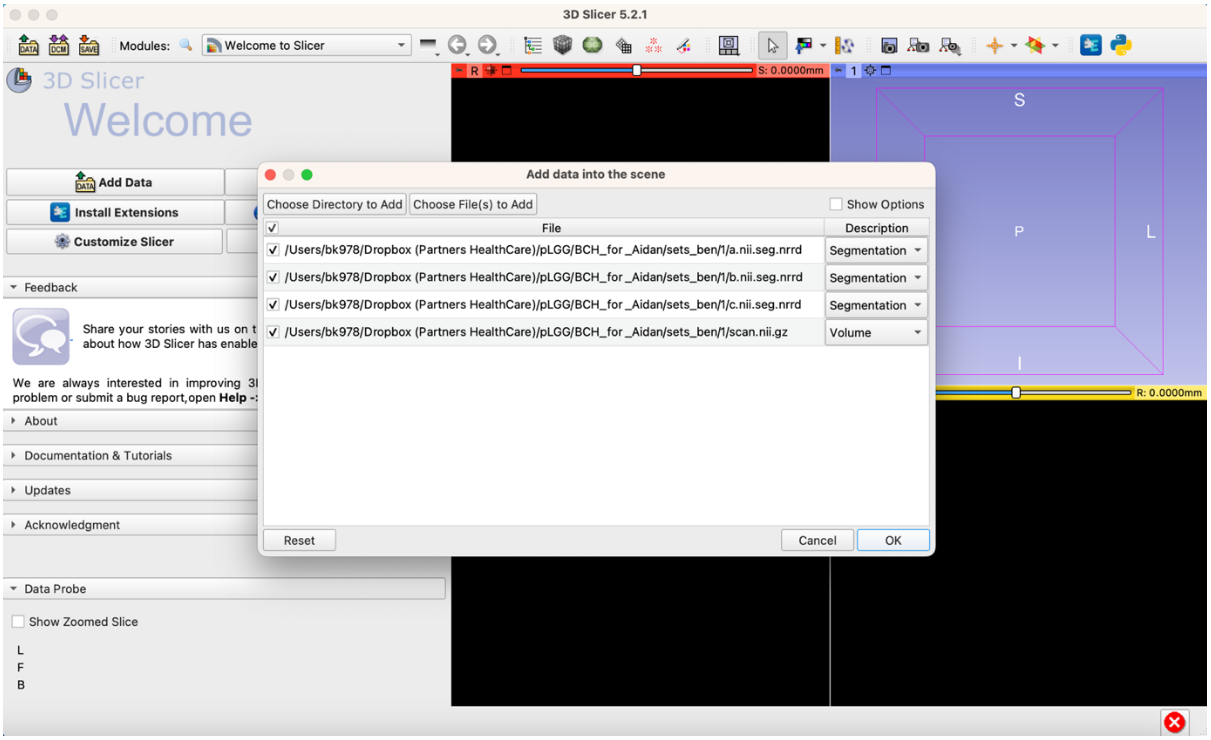


(3 Segmentations, 1 Volume)

4) Click "OK" and you should see three overlaid segmentations: (A: RED, B: YELLOW, C: BLUE)

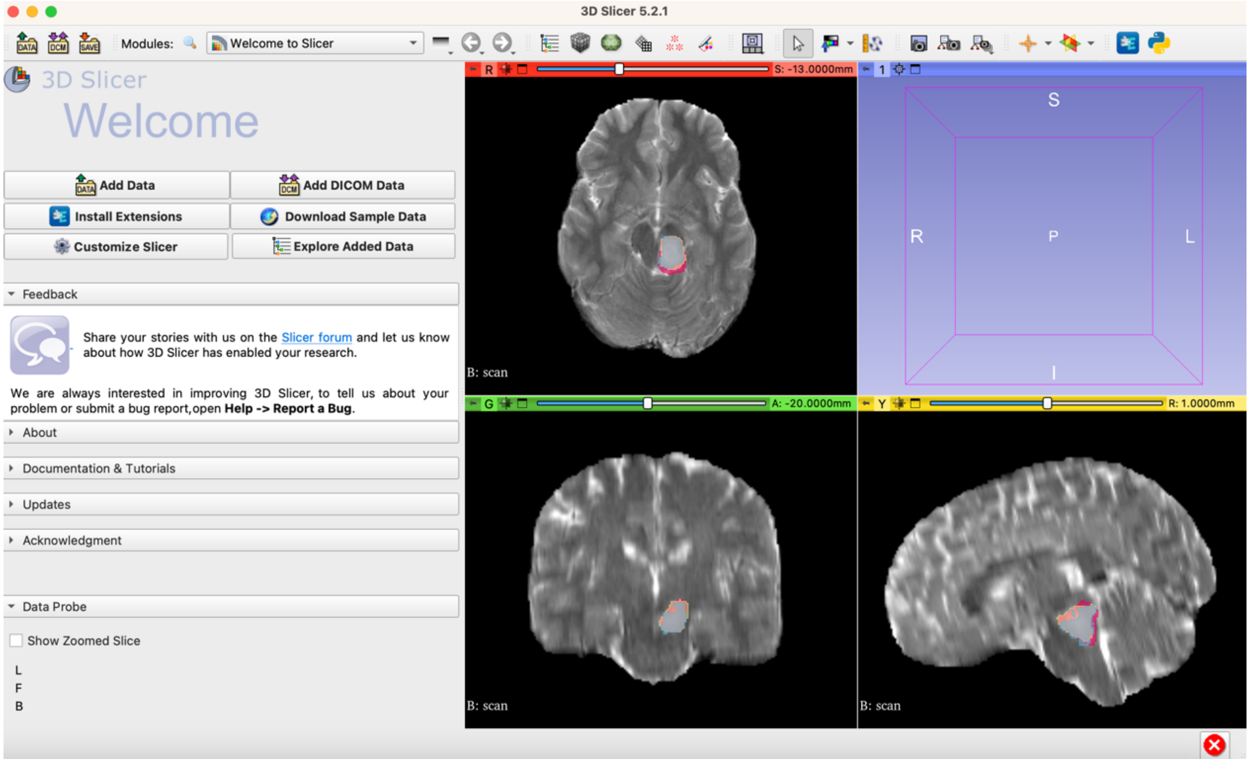


Use mouse to scroll, pan, and zoom as needed. Evaluate the three segmentations and complete the spreadsheet row.

Click File > "Close Scene" or Ctrl (or Cmd) +W to close the scene. Repeat steps for each scan.

Note: to toggle between semi-opaque segmentation and transparent outline, you can toggle the opacity by selecting the ">>" icon at the top left of the Red/Green/Yellow top window bar, and then selecting the following icon for each segmentation, one at a time…

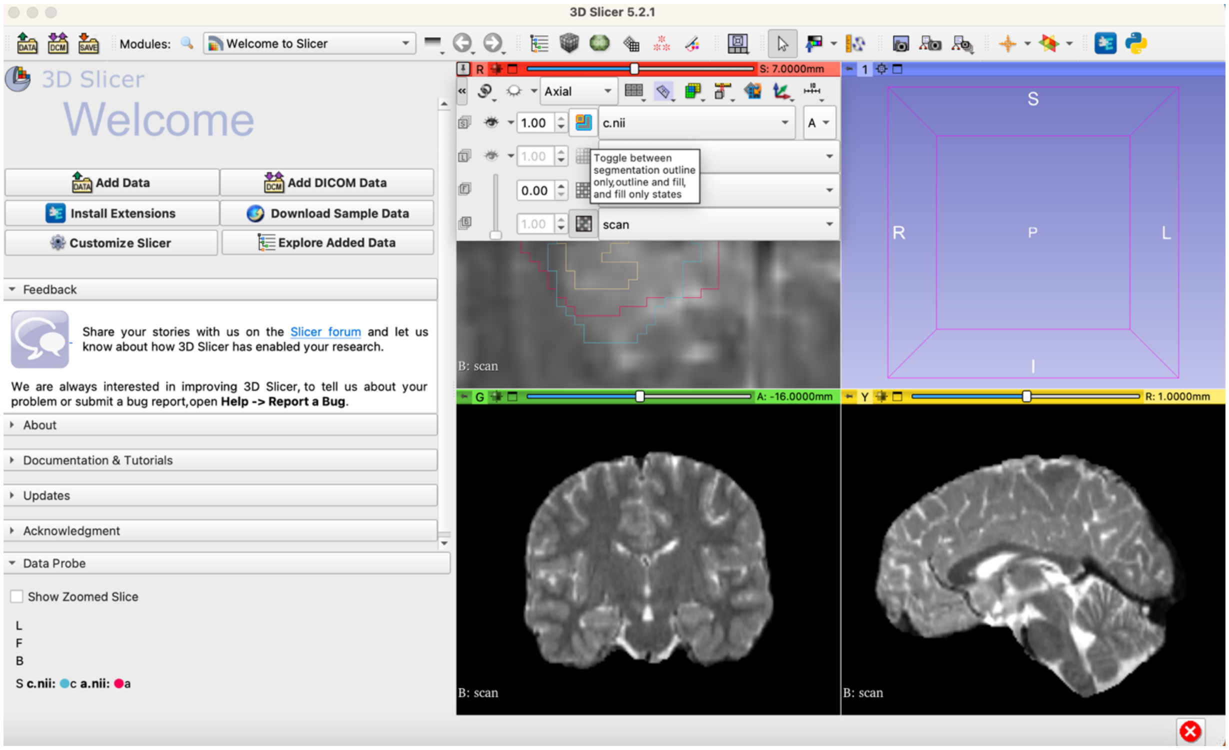
